# Blood glucose dynamics vary with economic provenance of meal

**DOI:** 10.1101/2024.11.21.24317693

**Authors:** Daniel Hamill, Trenton G. Smith, Bernard Venn

**Affiliations:** New Zealand Institute of Economic Research; Department of Economics, University of Otago; Department of Human Nutrition, University of Otago (retired)

## Abstract

The dynamics of blood sugar response to ultra-processed foods have strong parallels to the effects of addictive drugs. We hypothesize that if glycemic response is indeed an important determinant of habit formation–and hence product demand–then the largest producers of proprietary commercial foods will have formulated their products accordingly. We continuous glucose monitor (CGM) data from free-feeding adults spanning 579 meal events yielding a pooled time series dataset with more than 24,000 observations. We find strong evidence that addiction-like dynamic properties of the glycemic response (dose, rate of absorption, and withdrawal) are greater for globally branded fast food meals as compared to freshly-prepared or off-brand processed food meals. These differences are largely maintained when controlling for size of meal and nutrient content, suggesting that standard food labels may be insufficient to resolve what appears to be an important asymmetric information problem.

## Introduction

The recent epidemics of obesity and diabetes around the world have led public health advocates to call for general improvement in quality of diet, and left governments searching for appropriate public policy responses [1]. One of the difficulties in formulating food policies aimed at improving dietary quality is identifying specific product characteristics that i) can credibly be said to be exacerbating the targeted public health problems, and ii) can feasibly be regulated, for instance by mandating new labeling requirements. In this study we focus on one particular characteristic–glycemic response–with both intriguing behavioral characteristics and demonstrably negative health effects. Glycemic response (i.e., the increase in blood sugar [glucose] after eating a carbohydrate-rich food) has been noted, for example, to be particularly strong in industrially processed foods [2], and the dynamics of the glycemic response have been shown to mimic those of powerful drugs of addiction [3–6], suggesting that glycemic effects have the potential to be a powerful driver of product demand [5–9].

It has long been known that food processing can significantly impact glycemic response, independent of macronutrient content [10]. Our central hypothesis is that if high-glycemic food products are more likely to stimulate habit formation in consumers, then we should expect firms to invest in the discovery of product formulations that optimize this characteristic. Moreover, because both economies of scale and proprietary branding lend incentive to invest in product development, we hypothesize that globally branded fast food restaurant meals will have the strongest glycemic effects.

Our study is the first of its kind to examine fast food restaurant meals *as chosen and consumed by ordinary consumers*. This is important because it allows for variation along a crucial dimension: meal size. Fast food chains have many levers by which they can influence meal size, including advertisements, pricing scheme, product formulation (including proprietary flavor chemicals but also processing technologies and the use of salt/sweet/fat/protein to stimulate appetite), and even the lighting, signage, and aroma the consumer experiences when entering the restaurant. These strategies are often difficult for researchers to measure–and moreover are typically treated as trade secrets by the industry–but we can observe the end result: a meal is consumed and blood glucose rises.

Our experimental design makes use of continuous glucose monitoring (CGM) technology, which measures interstitial glucose every five minutes, 24 hours per day. We monitor the dietary behaviour of 12 non-diabetic adult subjects under free-feeding conditions for several days, yielding a large pooled time series dataset comprising 579 meal events and 24,084 individual observations. We find strong evidence that economic provenance of the meal is a strong predictor of the magnitude and dynamics of blood sugar perturbations. We also provide evidence that standard food labels provide insufficient information about glycemic response, as controlling for nutrient content attenuates but does not eliminate the effects we observe.

## Materials and methods

### Experimental Design

Twelve non-diabetic adult (age 18-65) volunteer subjects were recruited to participate in the study (written informed consent obtained, March 1 2016–April 30 2016). Each subject was asked to wear a Dexcom G4 Platinum CGM device [11] for 7 days, to record every ‘consumption event’ (meal or caloric beverage) with time-stamped photographs, and to consume at least one meal from a globally branded fast food restaurant over the course of their participation in the study. The Dexcom G4 measures interstitial glucose via a subcutaneous platinum probe, and must be calibrated by the user (via a finger stick test) twice per day. No other restrictions were placed on diet or behavior.

Photographic meal diaries were analyzed for nutritional content by a technician trained in the use of FoodWorks 8 software (Xyris Software Pty Ltd., Brisbane, Queensland, Australia), with verbal clarifications obtained from subjects where photographic evidence was ambiguous. FoodWorks 8 is widely used by dietitians and nutrition researchers to tabulate nutrient values for common food items or ingredients. In our regression analyses (see Regression Analysis below) we restrict our attention to the vector of nutrients available on the standard food label in New Zealand: calories, total carbohydrate, sugar, fiber, total fat, saturated fat, protein, sodium, and alcohol.^1^ The technician also categorized each meal as either freshly prepared, globally branded fast food, or other off-brand processed food.

### Data

Summary statistics for interstitial glucose by subject are shown in Table 1. Sample size per subject varied due to occasional gaps in the data (e.g., because the receiver was not in close proximity to the subject for a period of time), varying durations of CGM use, or (in the case of subject 8) because the transceiver battery failed before the study period ended. For the pooled sample of 24,084 observations mean interstitial glucose is 6.09 mmol/L, with standard deviation of 1.23.

**Table 1.** CGM Observations (mmol/L) by Subject.

| Subject | Mean | Std. Dev. | *N* |
| --- | --- | --- | --- |
| 1 | 6.52 | 1.44 | 2408 |
| 2 | 5.30 | 1.06 | 2249 |
| 3 | 5.64 | 0.96 | 1989 |
| 4 | 6.14 | 1.01 | 1925 |
| 5 | 6.91 | 1.02 | 1987 |
| 6 | 5.95 | 1.19 | 1930 |
| 7 | 5.56 | 1.05 | 1974 |
| 8 | 6.06 | 1.19 | 1392 |
| 9 | 6.16 | 0.71 | 1867 |
| 10 | 6.85 | 1.26 | 1918 |
| 11 | 6.38 | 1.40 | 2491 |
| 12 | 5.73 | 0.85 | 1954 |
| All | 6.09 | 1.23 | 24,084 |

Summary statistics for the 579 meals in our pooled sample are shown in Table 2, by meal type. There is clearly a high variance in meal size and nutrient content across all categories, but globally branded meals clearly dominate in every category, perhaps because many smaller meals or snacks are included in the others.

**Table 2.** Meal Characteristics.

| Nutrient | Meal Type |  |  |
| --- | --- | --- | --- |
|  | Fresh | Off-Brand | Global |
| Kilocalories | 234<br>(193) | 182<br>(220) | 916<br>(651) |
| Carbohydrate (g) | 25.0<br>(20.5) | 21.9<br>(30.2) | 103.6<br>(70.0) |
| Sugar (g) | 8.4<br>(10.6) | 10.0<br>(17.2) | 45.2<br>(48.8) |
| Fiber (g) | 3.1<br>(3.6) | 1.9<br>(3.9) | 5.9<br>(5.6) |
| Total Fat (g) | 9.6<br>(11.2) | 6.4<br>(9.3) | 35.9<br>(35.3) |
| Sat. Fat (g) | 3.8<br>(4.7) | 3.0<br>(4.6) | 13.3<br>(12.6) |
| Protein (g) | 10.8<br>(12.0) | 5.4<br>(7.7) | 41.6<br>(35.5) |
| Sodium (mg) | 314<br>(391) | 178<br>(358) | 1358<br>(1145) |
| Alcohol (g) | 0.35<br>(2.1) | 1.51<br>(4.6) | 0.44<br>(2.3) |
| *N* | 330 | 222 | 27 |

Note: Mean values shown (standard deviations in parentheses).

Our primary outcomes of interest are pre- and post-prandial blood glucose dynamics around meal events.^2^ Because these effects can extend for hours before and after a meal, and because our study specifies free-feeding conditions, it is naturally the case that many responses overlap with subsequent meals within our data. In order to allow for estimation of the effects of specific meal events, we create a vector of leading and lagged meal descriptor variables for 12 periods (60 minutes) before and 42 periods (210 minutes) after each meal. This vector includes binary indicators of meal type (Fresh, Off-Brand, Global Brand) as well as quantitative estimates of nutrient content for each nutrient in the meal.

### Regression Analysis

The usual concerns when analysing dynamic effects within a large time series sample such as ours are non-stationarity and serial correlation [12]. Because blood glucose regulation is known to be a homeostatic process, stationarity (e.g., no long-term time trends, no long-term temporal correlation between the dependent variable and independent variables) can safely be assumed. But our blood glucose data also certainly reflect unobserved shocks (e.g., due to physical exercise) that may persist over short periods of time, so we can expect serial correlation in our residuals–resulting in biased estimates of standard errors–if this problem is not addressed directly. We therefore implement a maximum likelihood regression model of the form:

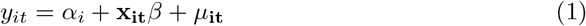

where *y*_*it*_ is interstitial glucose of subject *i* at time *t, α*_*i*_ is an individual-specific fixed effect, **x**_**it**_ is a vector of explanatory variables, and *μ*_*it*_ is an ARMA(*p, q*) disturbance term such that:

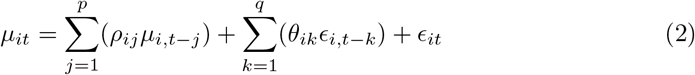

where *c*_*it*_ is a white-noise disturbance term. Exploratory analysis of the residuals from our main regression (Model 2 in Tables 3 and 4) suggests that the values *p* = 2 and *q* = 7 are sufficient to address serial correlation.^3,4^

**Table 3.** Dynamic Effects of Meals on Interstitial Glucose.

| Time Since<br>Meal (min) | Model 1: No Nutrient Controls |  |  | Model 2: Nutrient Controls |  |  |
| --- | --- | --- | --- | --- | --- | --- |
|  | Fresh | Off-Brand | Global | Fresh | Off-Brand | Global |
| -60 | -0.0146<br>(0.0114) | 0.00123<br>(0.0122) | -0.00225<br>(0.0482) | -0.00386<br>(0.0110) | 0.0187<br>(0.0129) | 0.0382<br>(0.0410) |
| -55 | -0.0293<br>(0.0211) | 0.0126<br>(0.0218) | -0.0363<br>(0.0950) | -0.0137<br>(0.0230) | 0.0372<br>(0.0288) | 0.0461<br>(0.110) |
| -50 | -0.0287<br>(0.0291) | 0.00355<br>(0.0225) | -0.102<br>(0.110) | -0.0141<br>(0.0347) | 0.0267<br>(0.0354) | 0.0185<br>(0.114) |
| -45 | -0.0280<br>(0.0359) | -0.00632<br>(0.0311) | -0.145<br>(0.126) | -0.00262<br>(0.0424) | 0.0330<br>(0.0504) | 0.0460<br>(0.118) |
| -40 | -0.0350<br>(0.0369) | -0.00910<br>(0.0464) | -0.164<br>(0.124) | -0.000861<br>(0.0413) | 0.0340<br>(0.0653) | 0.0746<br>(0.117) |
| -35 | -0.0454<br>(0.0410) | -0.00993<br>(0.0509) | -0.207<br>(0.135) | 0.00279<br>(0.0491) | 0.0395<br>(0.0684) | 0.109<br>(0.137) |
| -30 | -0.0682*<br>(0.0377) | -0.0288<br>(0.0532) | -0.221<br>(0.144) | -0.0122<br>(0.0433) | 0.0313<br>(0.0727) | 0.132<br>(0.149) |
| -25 | -0.0842**<br>(0.0400) | -0.0687<br>(0.0615) | -0.248*<br>(0.137) | -0.0214<br>(0.0468) | 0.00332<br>(0.0767) | 0.130<br>(0.147) |
| -20 | -0.0852**<br>(0.0412) | -0.0884<br>(0.0632) | -0.274*<br>(0.142) | -0.0219<br>(0.0504) | -0.00456<br>(0.0762) | 0.0853<br>(0.172) |
| -15 | -0.0836**<br>(0.0394) | -0.0838<br>(0.0582) | -0.310**<br>(0.152) | -0.0169<br>(0.0535) | 0.0122<br>(0.0660) | 0.0862<br>(0.187) |
| -10 | -0.0882**<br>(0.0343) | -0.0784<br>(0.0562) | -0.320**<br>(0.160) | -0.0275<br>(0.0519) | 0.0213<br>(0.0542) | 0.0596<br>(0.192) |
| -5 | -0.0816**<br>(0.0360) | -0.0786<br>(0.0554) | -0.315**<br>(0.140) | -0.0193<br>(0.0471) | 0.0147<br>(0.0513) | 0.0813<br>(0.166) |
| 0 | -0.0784**<br>(0.0392) | -0.0939*<br>(0.0526) | -0.305*<br>(0.161) | -0.0135<br>(0.0447) | -0.00283<br>(0.0486) | 0.0693<br>(0.178) |
| 5 | -0.0871**<br>(0.0444) | -0.128**<br>(0.0553) | -0.267<br>(0.194) | -0.00857<br>(0.0406) | -0.0370<br>(0.0475) | 0.147<br>(0.180) |
| 10 | -0.0791<br>(0.0490) | -0.137**<br>(0.0610) | -0.199<br>(0.227) | 0.00479<br>(0.0411) | -0.0410<br>(0.0532) | 0.214<br>(0.183) |
| 15 | -0.0251<br>(0.0510) | -0.129*<br>(0.0672) | -0.0685<br>(0.257) | 0.0742*<br>(0.0426) | -0.0206<br>(0.0640) | 0.374**<br>(0.185) |
| 20 | 0.0679<br>(0.0474) | -0.0556<br>(0.0703) | 0.0308<br>(0.250) | 0.147***<br>(0.0362) | 0.0424<br>(0.0711) | 0.417**<br>(0.170) |
| 25 | 0.177***<br>(0.0450) | 0.0645<br>(0.0686) | 0.164<br>(0.248) | 0.243***<br>(0.0337) | 0.161**<br>(0.0730) | 0.518***<br>(0.185) |
| 30 | 0.321***<br>(0.0563) | 0.238***<br>(0.0647) | 0.391<br>(0.250) | 0.353***<br>(0.0373) | 0.304***<br>(0.0693) | 0.611***<br>(0.178) |
| 35 | 0.460***<br>(0.0824) | 0.401***<br>(0.0786) | 0.643**<br>(0.251) | 0.444***<br>(0.0668) | 0.427***<br>(0.0875) | 0.710***<br>(0.195) |

Table 3 (continued): Dynamic Effects of Meals on Interstitial Glucose
| Time Since<br>Meal (min) | Model 1: No Nutrient Controls |  |  | Model 2: Nutrient Controls |  |  |
| --- | --- | --- | --- | --- | --- | --- |
|  | Fresh | Off-Brand | Global | Fresh | Off-Brand | Global |
| 40 | 0.571***<br>(0.115) | 0.514***<br>(0.0966) | 0.794***<br>(0.275) | 0.504***<br>(0.105) | 0.487***<br>(0.107) | 0.650**<br>(0.259) |
| 45 | 0.606***<br>(0.144) | 0.555***<br>(0.107) | 0.997***<br>(0.306) | 0.496***<br>(0.134) | 0.483***<br>(0.115) | 0.678**<br>(0.287) |
| 50 | 0.601***<br>(0.161) | 0.545***<br>(0.115) | 1.170***<br>(0.339) | 0.471***<br>(0.151) | 0.429***<br>(0.121) | 0.743**<br>(0.303) |
| 55 | 0.555***<br>(0.156) | 0.479***<br>(0.123) | 1.300***<br>(0.338) | 0.421***<br>(0.144) | 0.342***<br>(0.122) | 0.819***<br>(0.295) |
| 60 | 0.487***<br>(0.148) | 0.377***<br>(0.118) | 1.376***<br>(0.350) | 0.377***<br>(0.144) | 0.235**<br>(0.116) | 0.873***<br>(0.320) |
| 65 | 0.430***<br>(0.130) | 0.288***<br>(0.110) | 1.472***<br>(0.371) | 0.347**<br>(0.137) | 0.168<br>(0.107) | 1.034***<br>(0.366) |
| 70 | 0.378***<br>(0.110) | 0.218**<br>(0.107) | 1.544***<br>(0.402) | 0.332**<br>(0.135) | 0.135<br>(0.102) | 1.202***<br>(0.444) |
| 75 | 0.338***<br>(0.0928) | 0.170<br>(0.106) | 1.607***<br>(0.405) | 0.312**<br>(0.135) | 0.109<br>(0.104) | 1.307**<br>(0.510) |
| 80 | 0.304***<br>(0.0797) | 0.149<br>(0.103) | 1.613***<br>(0.415) | 0.289**<br>(0.125) | 0.103<br>(0.105) | 1.309**<br>(0.556) |
| 85 | 0.280***<br>(0.0655) | 0.118<br>(0.0996) | 1.514***<br>(0.408) | 0.261**<br>(0.115) | 0.0786<br>(0.100) | 1.194**<br>(0.579) |
| 90 | 0.264***<br>(0.0587) | 0.110<br>(0.0952) | 1.427***<br>(0.395) | 0.238**<br>(0.104) | 0.0702<br>(0.101) | 1.133**<br>(0.576) |
| 95 | 0.245***<br>(0.0626) | 0.103<br>(0.0877) | 1.340***<br>(0.356) | 0.220**<br>(0.0957) | 0.0583<br>(0.0958) | 1.044**<br>(0.519) |
| 100 | 0.239***<br>(0.0662) | 0.0887<br>(0.0793) | 1.189***<br>(0.318) | 0.217**<br>(0.0853) | 0.0487<br>(0.0890) | 0.904**<br>(0.447) |
| 105 | 0.214***<br>(0.0693) | 0.0787<br>(0.0732) | 1.044***<br>(0.285) | 0.186**<br>(0.0814) | 0.0389<br>(0.0802) | 0.787**<br>(0.386) |
| 110 | 0.195***<br>(0.0707) | 0.0689<br>(0.0682) | 0.994***<br>(0.237) | 0.176**<br>(0.0817) | 0.0432<br>(0.0745) | 0.799**<br>(0.332) |
| 115 | 0.190***<br>(0.0660) | 0.0549<br>(0.0657) | 0.961***<br>(0.226) | 0.164**<br>(0.0720) | 0.0263<br>(0.0703) | 0.748**<br>(0.310) |
| 120 | 0.196***<br>(0.0592) | 0.0579<br>(0.0623) | 0.995***<br>(0.209) | 0.150**<br>(0.0624) | 0.0238<br>(0.0703) | 0.774**<br>(0.322) |
| 125 | 0.199***<br>(0.0527) | 0.0445<br>(0.0537) | 0.952***<br>(0.210) | 0.141***<br>(0.0543) | 0.00994<br>(0.0653) | 0.711**<br>(0.313) |
| 130 | 0.205***<br>(0.0510) | 0.0266<br>(0.0560) | 0.967***<br>(0.216) | 0.138***<br>(0.0509) | -0.00375<br>(0.0678) | 0.712**<br>(0.329) |
| 135 | 0.220***<br>(0.0441) | 0.0216<br>(0.0590) | 0.952***<br>(0.211) | 0.158***<br>(0.0464) | -0.00602<br>(0.0714) | 0.750**<br>(0.336) |

Table 3 (continued): Dynamic Effects of Meals on Interstitial Glucose
| Time Since<br>Meal (min) | Model 1: No Nutrient Controls |  |  | Model 2: Nutrient Controls |  |  |
| --- | --- | --- | --- | --- | --- | --- |
|  | Fresh | Off-Brand | Global | Fresh | Off-Brand | Global |
| 140 | 0.199***<br>(0.0457) | 0.0207<br>(0.0616) | 0.903***<br>(0.218) | 0.136***<br>(0.0510) | -0.0149<br>(0.0672) | 0.698**<br>(0.338) |
| 145 | 0.174***<br>(0.0519) | 0.0352<br>(0.0577) | 0.858***<br>(0.228) | 0.118**<br>(0.0551) | 0.0122<br>(0.0654) | 0.677*<br>(0.354) |
| 150 | 0.151***<br>(0.0552) | 0.0319<br>(0.0454) | 0.812***<br>(0.238) | 0.0959<br>(0.0632) | 0.0181<br>(0.0521) | 0.654*<br>(0.369) |
| 155 | 0.135**<br>(0.0543) | 0.0282<br>(0.0325) | 0.762***<br>(0.244) | 0.0768<br>(0.0642) | 0.0181<br>(0.0415) | 0.589<br>(0.381) |
| 160 | 0.113**<br>(0.0541) | 0.00372<br>(0.0292) | 0.746***<br>(0.259) | 0.0612<br>(0.0682) | 0.00170<br>(0.0414) | 0.572<br>(0.393) |
| 165 | 0.0875<br>(0.0536) | -0.0207<br>(0.0354) | 0.735***<br>(0.243) | 0.0391<br>(0.0787) | -0.0128<br>(0.0464) | 0.572<br>(0.353) |
| 170 | 0.0654<br>(0.0454) | -0.0305<br>(0.0444) | 0.708***<br>(0.219) | 0.0104<br>(0.0739) | -0.0312<br>(0.0527) | 0.515*<br>(0.277) |
| 175 | 0.0332<br>(0.0403) | -0.0421<br>(0.0500) | 0.679***<br>(0.213) | -0.0286<br>(0.0710) | -0.0464<br>(0.0573) | 0.430**<br>(0.215) |
| 180 | 0.0243<br>(0.0293) | -0.0454<br>(0.0505) | 0.619***<br>(0.218) | -0.0529<br>(0.0623) | -0.0629<br>(0.0568) | 0.309*<br>(0.187) |
| 185 | 0.0262<br>(0.0223) | -0.0190<br>(0.0482) | 0.532**<br>(0.234) | -0.0473<br>(0.0530) | -0.0362<br>(0.0516) | 0.235<br>(0.199) |
| 190 | 0.0240<br>(0.0174) | 0.00863<br>(0.0452) | 0.413*<br>(0.229) | -0.0432<br>(0.0395) | -0.00838<br>(0.0441) | 0.133<br>(0.207) |
| 195 | 0.0233<br>(0.0178) | 0.0167<br>(0.0440) | 0.312*<br>(0.188) | -0.0405<br>(0.0304) | -0.00442<br>(0.0422) | 0.0636<br>(0.160) |
| 200 | 0.0178<br>(0.0208) | 0.0139<br>(0.0341) | 0.171<br>(0.123) | -0.0268<br>(0.0220) | -0.00171<br>(0.0358) | -0.0316<br>(0.107) |
| 205 | 0.00855<br>(0.0158) | -0.00421<br>(0.0243) | 0.0546<br>(0.0793) | -0.0133<br>(0.0148) | -0.0137<br>(0.0250) | -0.0399<br>(0.0711) |
| 210 | -0.00171<br>(0.00839) | 0.00330<br>(0.0141) | 0.00151<br>(0.0343) | -0.00890<br>(0.00956) | 0.000507<br>(0.0151) | -0.0241<br>(0.0387) |
| *N* | 24,084 |  |  | 24,084 |  |  |

Notes: Semi-robust standard errors (adjusted for clustering by individual) in parentheses. Both models include individual fixed effects and AR(2)/MA(8) error structure. Marginal effects of individual nutrients in Model 2 are reported in Table 4.
\*\*\* $p < 0.01$ , \*\* $p < 0.05$ , \* $p < 0.1$

**Table 4.**
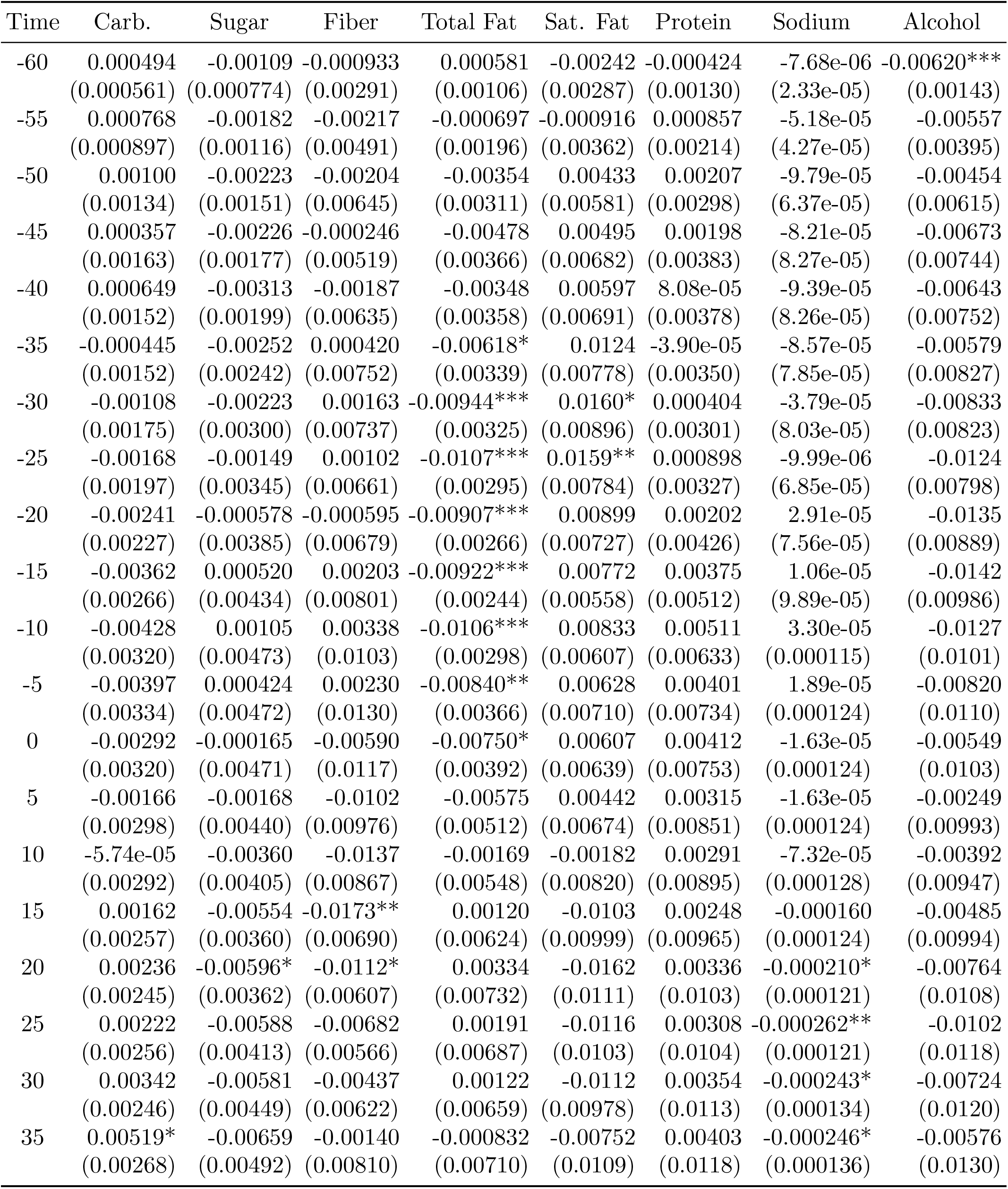
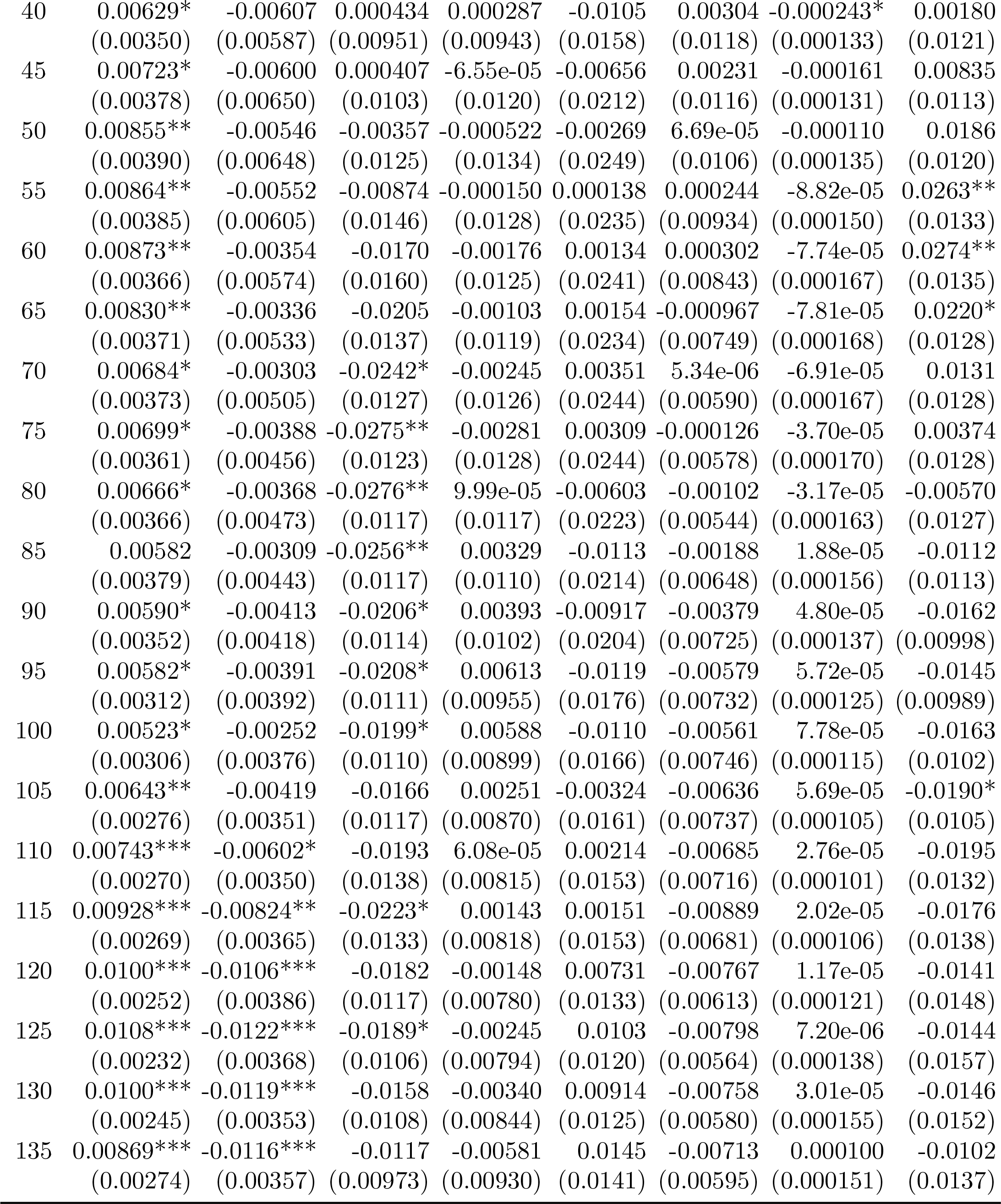
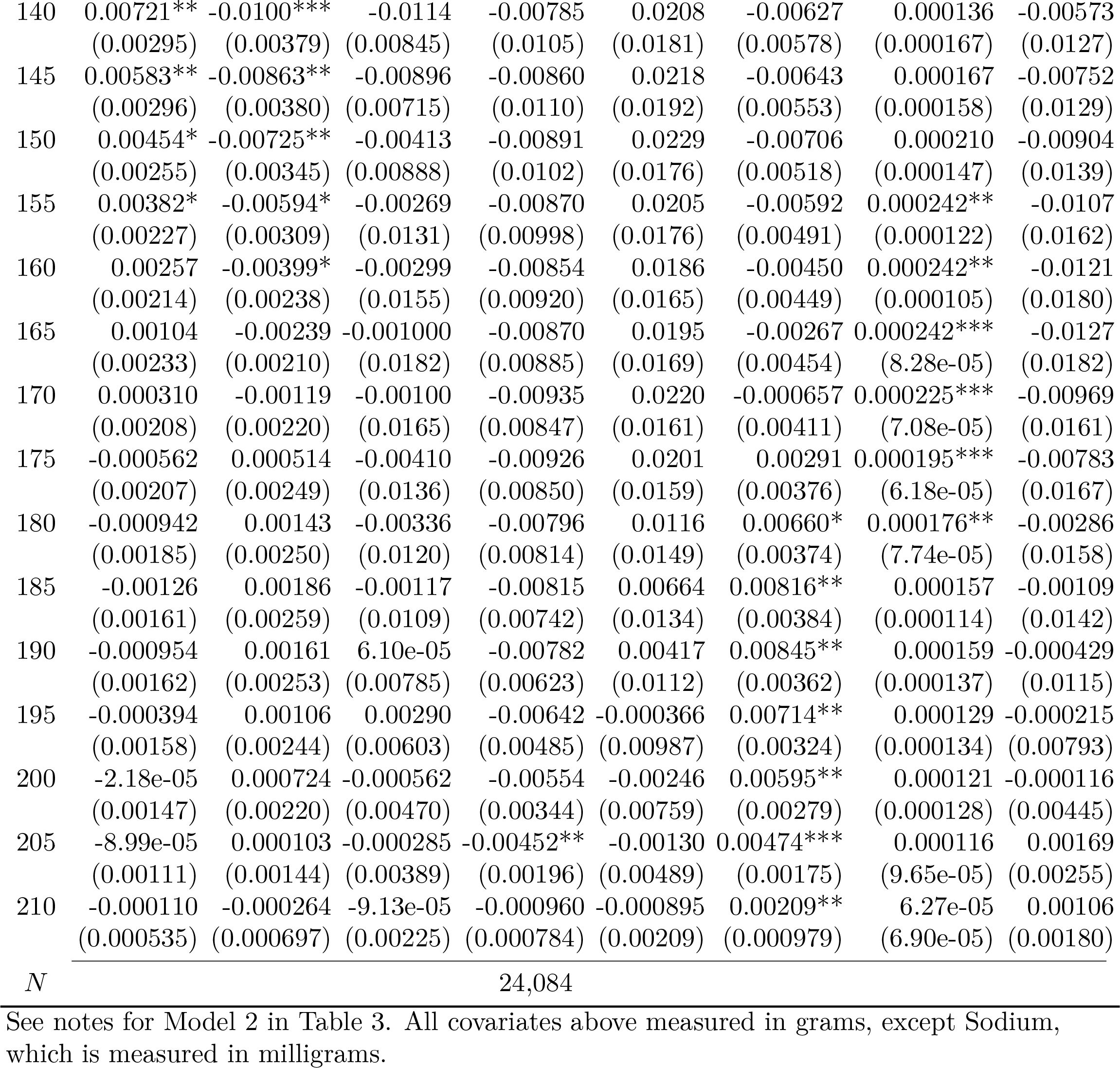
Dynamic Effects of Meal Nutrient Content on Interstitial Glucose.

## Results

Our main results are presented in Table 3 (and represented graphically in Figures 1 and 2). In Model 1, no controls for meal size or nutrient content are included, yielding estimates of blood sugar response by meal type (Fig 1). For all three meal types, the largest estimates are significantly different from zero (Table 3), with peak interstitial glucose occurring at 45 minutes for Fresh and Off-Brand meals, and 80 minutes for Global Brand meals. We also use these coefficient estimates to construct four alternative measures of addiction dynamics: “craving” (*C*), the area above the curve in the pre-meal period (effectively, the sum of negative coefficients in a given column of Table 3) “dose” (*D*), the area under the curve in Figure 1 (effectively, the sum of positive coefficients in a given column of Table 3); “rate of absorption” (*A*), the maximal increase in interstitial glucose over any 40-minute period; and “withdrawal” (*W*), the maximal decrease in interstitial glucose over any 40-minute period. We provide the results of hypothesis tests on the magnitudes of each of these measures in Table 5, both against a null hypothesis of zero and in pairwise tests of equality by meal type. We find that every measure but one (craving for Off-Brand, p=0.0809) is significantly greater than zero at 5% significance for all three meal types. In pairwise comparisons, both dose and rate of absorption are significantly greater for Global Brand than for either Fresh or Off-Brand, but Fresh and Off-Brand are not significantly different. None of the pairwise comparisons for craving or withdrawal are significant at *α* = 0.05.

**Table 5.**
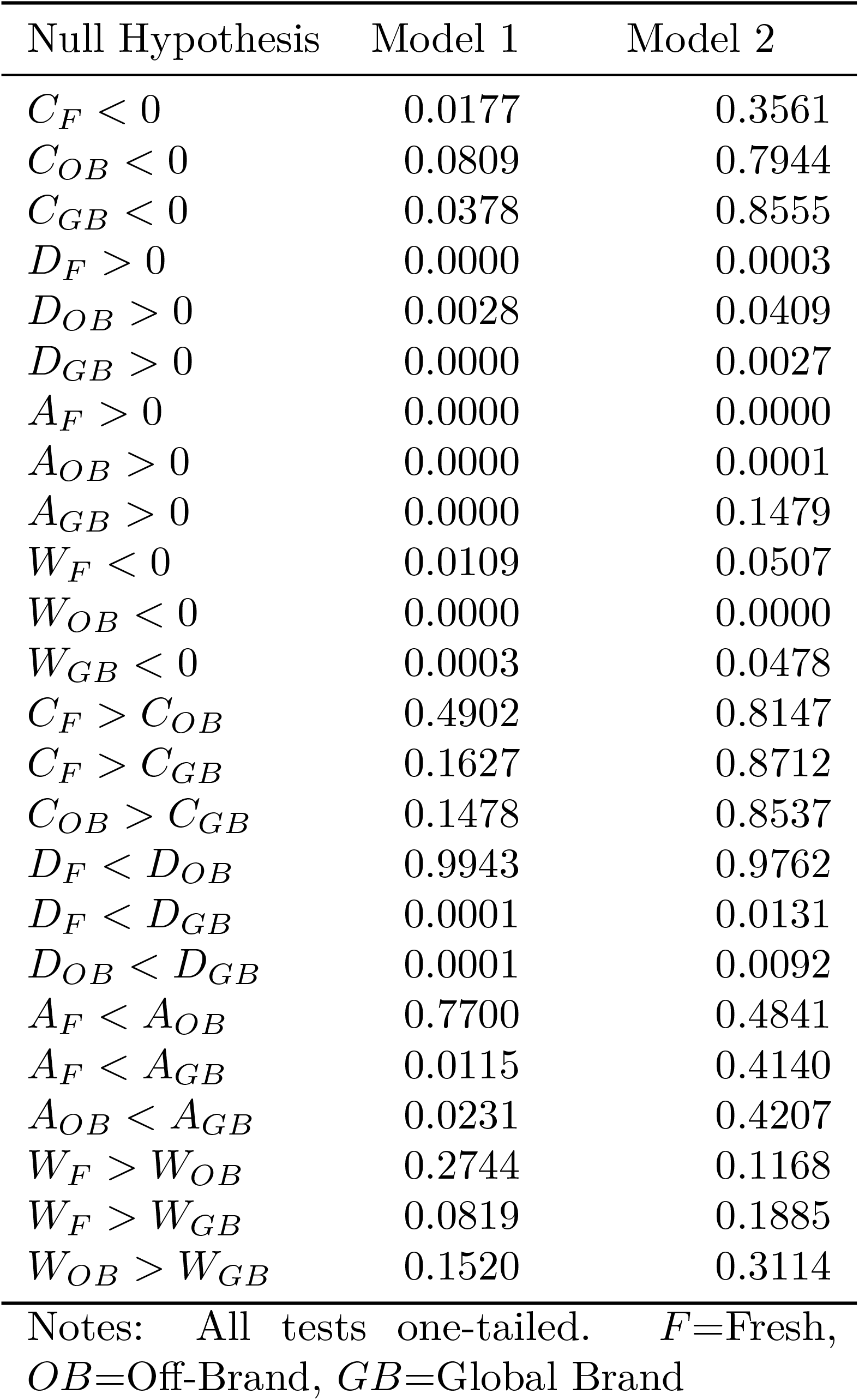
Chi-Squared Tests (*p*-values) on Measures of Glycemic Dose (*D*), Rate of Absorption (*A*), and Withdrawal (*W*)

| Null Hypothesis | Model 1 | Model 2 |
| --- | --- | --- |
| $C_F < 0$ | 0.0177 | 0.3561 |
| $C_{OB} < 0$ | 0.0809 | 0.7944 |
| $C_{GB} < 0$ | 0.0378 | 0.8555 |
| $D_F > 0$ | 0.0000 | 0.0003 |
| $D_{OB} > 0$ | 0.0028 | 0.0409 |
| $D_{GB} > 0$ | 0.0000 | 0.0027 |
| $A_F > 0$ | 0.0000 | 0.0000 |
| $A_{OB} > 0$ | 0.0000 | 0.0001 |
| $A_{GB} > 0$ | 0.0000 | 0.1479 |
| $W_F < 0$ | 0.0109 | 0.0507 |
| $W_{OB} < 0$ | 0.0000 | 0.0000 |
| $W_{GB} < 0$ | 0.0003 | 0.0478 |
| $C_F > C_{OB}$ | 0.4902 | 0.8147 |
| $C_F > C_{GB}$ | 0.1627 | 0.8712 |
| $C_{OB} > C_{GB}$ | 0.1478 | 0.8537 |
| $D_F < D_{OB}$ | 0.9943 | 0.9762 |
| $D_F < D_{GB}$ | 0.0001 | 0.0131 |
| $D_{OB} < D_{GB}$ | 0.0001 | 0.0092 |
| $A_F < A_{OB}$ | 0.7700 | 0.4841 |
| $A_F < A_{GB}$ | 0.0115 | 0.4140 |
| $A_{OB} < A_{GB}$ | 0.0231 | 0.4207 |
| $W_F > W_{OB}$ | 0.2744 | 0.1168 |
| $W_F > W_{GB}$ | 0.0819 | 0.1885 |
| $W_{OB} > W_{GB}$ | 0.1520 | 0.3114 |

Notes: All tests one-tailed. $F$ =Fresh, $OB$ =Off-Brand, $GB$ =Global Brand

**Figure 1:**
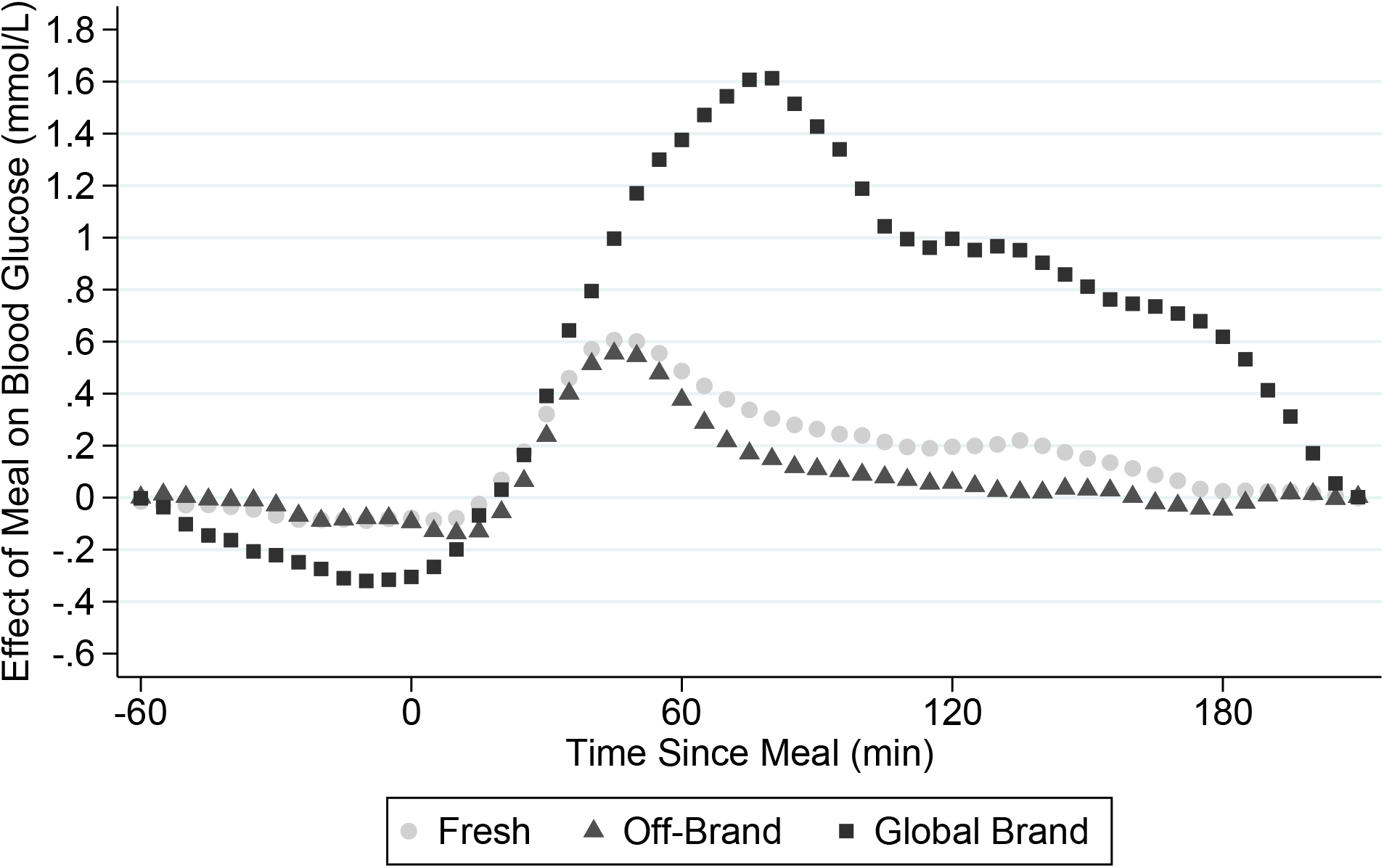
Blood Glucose Response by Meal Type (not controlling for nutrient content)

**Figure 2:**
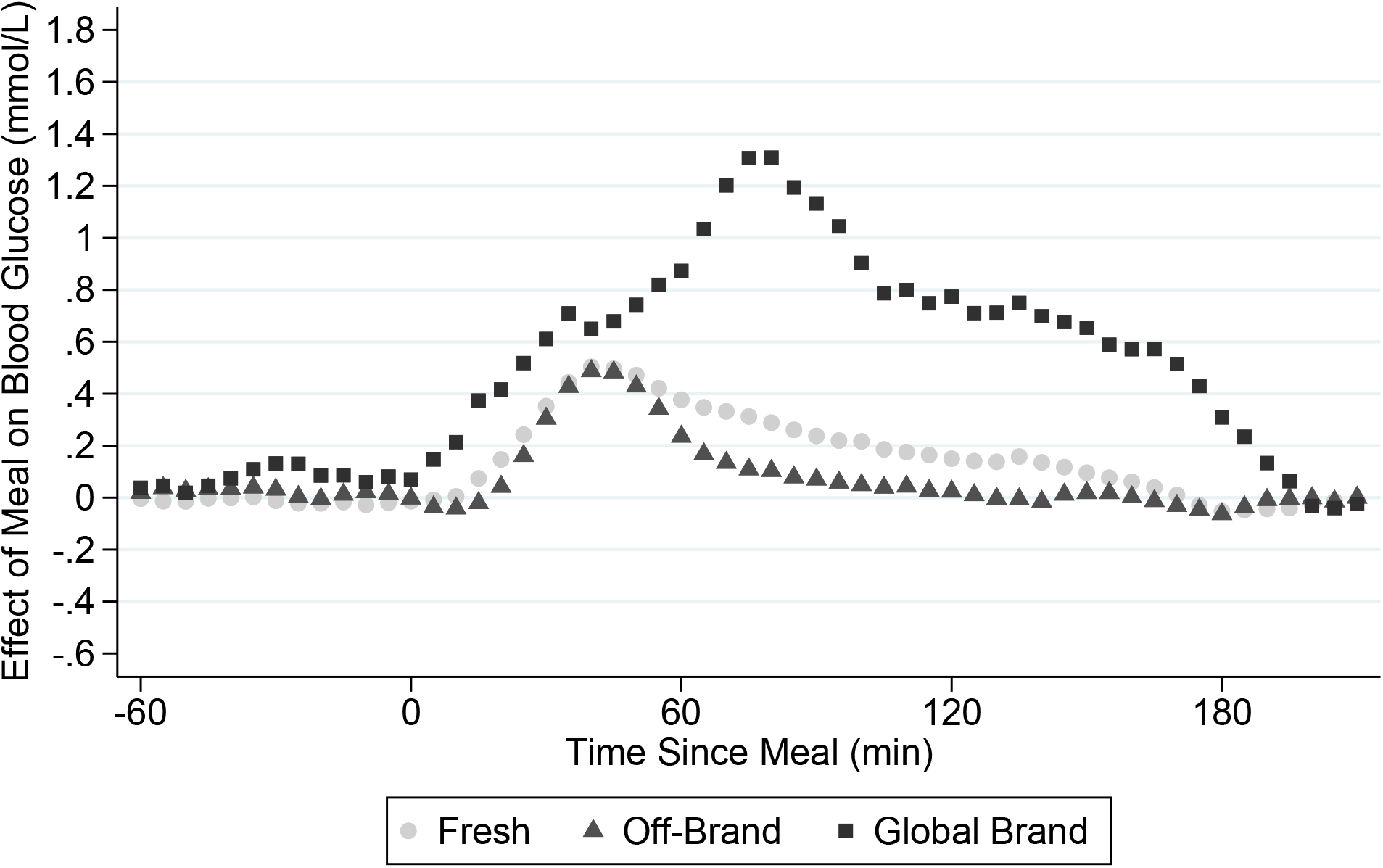
Blood Glucose Response by Meal Type (controlling for nutrient content)

Controlling for nutrients (Model 2 in Table 3, and Figure 2), the effects of meal type are attenuated but not eliminated. In principle, if the nutrient label provides sufficient information about glycemic effects (subject to several important caveats, discussed below), every coefficient shown for Model 2 in Table 3 should be indistinguishable from zero. The fact that many are not, and that several of the hypothesis tests on blood sugar dynamics (Table 5) are maintained in Model 2 is suggestive, at least, that current food labels provide insufficient information for consumers to be able to predict glycemic response based on the label alone. Indeed, our estimates allow us to quantify this deficiency. Focusing on dose (the most widely used measure of glycemic effects) and taking the coefficients in Model 1 to represent 100% of the blood sugar response by meal type, controlling for nutrients reduces our estimates of dose by only about 17% for Fresh, 18% for Off-Brand, and 20% for Global Brand.

Finally, we report the marginal effects of nutrients on glycemic response in Table 4. Predictably, carbohydrate has the largest and most statistically significant positive effect, with sugar (a component of carbohydrate that includes a large proportion of fructose, which is not detected by CGM) having a significant negative effect late in the postprandial period. We also observe statistically significant negative effects of fat in the preprandial period.

## Discussion

This study is the result of a cross-disciplinary collaboration, and this is reflected in our experimental design. Proprietary food products produced at scale are enormously profitable.^5^ There is every reason to expect that the largest players in the industry have invested heavily in optimizing their product in ways that stimulate demand [14].

Consumer researchers, meanwhile, have begun to use the term “food addiction,” and evidence from neuroscience strongly suggests this is not mere hyperbole [15]. For economists and the food regulators they advise, all of this would be quite harmless if we could be confident that consumers were fully informed about the consequences of their choices [16–18]. All of this leads us to a free-feeding experimental design, a focus on the economic provenance of meals and its relation to addiction-like dynamics, and statistical controls for available information (in the form of today’s mandatory nutrient label). While our findings are not definitive (see below), they are striking.

Several caveats should be mentioned regarding the results presented as Model 2 in Table 3, along with the corresponding hypothesis tests in Table 5. It is possible that these results could be explained not by deficiencies in the food label, but rather by i) measurement error in translating photo diaries to nutrients; ii) non-linear effects of individual nutrients; or iii) interaction effects between nutrients that impact the magnitude or timing of the glycemic response. But none of these explanations would seem to obviate the need for improved information on the label: if a trained (and computer-assisted) scientist cannot easily estimate nutrient amounts, or if complex non-linear transformations of nutrient amounts are required to tease out potential glycemic effects, then surely these effects will remain largely unknown to the typical consumer.

## Data Availability

Data cannot be shared publicly because of privacy restrictions imposed by the University of Otago Human Ethics Committee.

## Acknowledgments

The authors are grateful for helpful comments from colleagues in the departments of Economics and Human Nutrition; from seminar participants at Colorado State University, the University of Wyoming, Tufts University, Newcastle University, and the University of Bozen-Bolzano; and from participants at the 2017 Applied & Agricultural Economics Meetings and the 2018 Australia-New Zealand Workshop in Experimental Economics; and for the careful advice of the Human Ethics Committee (Health) at the University of Otago. All remaining errors are our own.

## Footnotes

1 In practice, calories is dropped from regressions due to perfect collinearity with the vector of macronutrients.

2 While our statistical analysis focuses on variation at the level of meals (rather than individuals) we note that in fully 50% of our subjects, the highest recorded blood glucose reading occured within 90 minutes of consumption of a globally branded fast food meal.

3 This was verified via a grid search over values of *p* and *q* of the autocorrelations and partial autocorrelations of residuals, as well as by the sequential Cumby-Huizinga ([13]) test with α = 0:05. We note further that these values are physiologically plausible.

4 In practice, Eq (1) was estimated using the arima command in Stata 14.

5 The market for globally branded fast foods generates annual revenues exceeding $1.1 trillion (https://www.ibisworld.com, accessed 27 October 2024).

